# Functional Assessment of Protein Variants in Structured Domains by Fluorescence Cross-Correlation Spectroscopy

**DOI:** 10.1101/2024.05.23.24307779

**Authors:** Àngels Mateu-Regué, Luca Mariani, Frederik Otzen Bagger, Muthiah Bose, Finn Cilius Nielsen

## Abstract

With the expanding catalogue of novel disease-genes, there is an increasing need to establish the significance of potential disease-causing variants. Based on the idea that pathogenic variants in structured protein domains disturb folding and association with macromolecular assemblies, we employed Fluorescence Correlation and Cross-Correlation Spectroscopy (FCS and FCCS) to assess *in vivo* protein complex formation. Since the molecular underpinning of BRCA-associated breast and ovarian cancers is well defined and data from a recent genome editing screening allowed us to compare variant binding data with a reliable functional HRD test in addition to ClinVar and AlphaMissense data, we examined the binding of mutated wild-type BRCA1 or isolated RING and BRCT domains to BARD1 and RBBP8, respectively. The results demonstrate that FCCS, whether applied to full-length BRCA1 in live cells and/or to isolated domains in cellular lysates identified pathogenic BRCA1 RING or BRCT domain variants. We moreover demonstrate the feasibility of employing FCCS for analysis of HNPCC-related factor MSH2 and MEN1 factor Menin variants in combination with DNA mismatch repair factor MSH6 and transcription factor JUND, respectively. We propose that FCCS may be an appealing complement to current clinical procedures for classifying variants, for many monogenic diseases given its generic nature and ease of use.

## INTRODUCTION

The rapid advancements in genomics have enhanced our comprehension of the genetic underpinnings of various rare diseases. With an ever-expanding catalogue of novel disease-associated genetic variations, there is a pressing need to establish the clinical significance of potential disease-causing variants. Traditionally, variant classification depended on the nature of the variant and its known association or co-segregation with a specific disease. However, co-segregation is not always feasible, leading to the contemporary utilization of ACMG/AMP guidelines^1^, which based on data from functional studies, segregation analyses, or clinical correlations categorize variants as pathogenic, likely pathogenic, variants of uncertain significance (VUS), likely benign, or benign. VUS, in particular, pose a significant clinical challenge as they leave patients in a state of uncertainty. To address this challenge, the development of large-scale variant databases, enhanced computational predictive tools like AlphaMissense^2^, and the advancement of functional analyses have represented significant steps forward. Two notable examples of functional analyses involve the early use of minigenes^3^ to investigate mutations affecting mRNA splicing, and more recently, systematic screening approaches such as the BRCA1 saturation editing screening^4^, aimed at assessing homologous recombination deficiency. However, there is currently no single, universally applicable approach for functional testing of protein variants, which impedes the application of functional analyses in the clinical environment.

Proteins are fundamental to cellular processes, and it is estimated that the majority of proteins within a given proteome exert their function as part of complexes with other factors^5^. Assembly is governed by conserved structured domains within the proteins and pathogenic variants frequently disrupt the structure of these domains, leading to misfolding of the proteins and destabilization of the functional units^6-9^. Various methods, such as affinity purification coupled with mass spectrometry (AP-MS), yeast two-hybrid (Y2H) assays, and computational approaches, have effectively been employed to characterize protein-protein complexes. While these methods have contributed significantly to our understanding of cellular protein networks, they are labour-intensive and may not always be readily adaptable for clinical analyses. Fluorescence Correlation and Cross-Correlation Spectroscopy (FCS and FCCS) offer alternative generic approaches to rapidly and precisely assess protein diffusion, stoichiometry, and complex formation (Figure 1) ^10,11^. FCS is based on the recording of fluorescence fluctuations produced by labelled molecules or proteins entering and exiting a small focal volume and analysing these fluctuations through autocorrelation. FCCS, on the other hand, leverages cross-correlation between molecules or proteins labelled with distinct spectral markers to determine if two factors associate or are part of the same macromolecular complex^10^. FCS and FCCS are versatile and have been applied to analysis of various cellular processes, such as, protein-protein interactions, protein-nucleic acid interactions, receptor-ligand binding, and molecular diffusion dynamics. They offer single-molecule sensitivity and can analyse proteins and their interactions both in live cells and *in vitro*. From a clinical perspective, they are also be appreciated for their speed and reproducibility.

**Figure 1.**
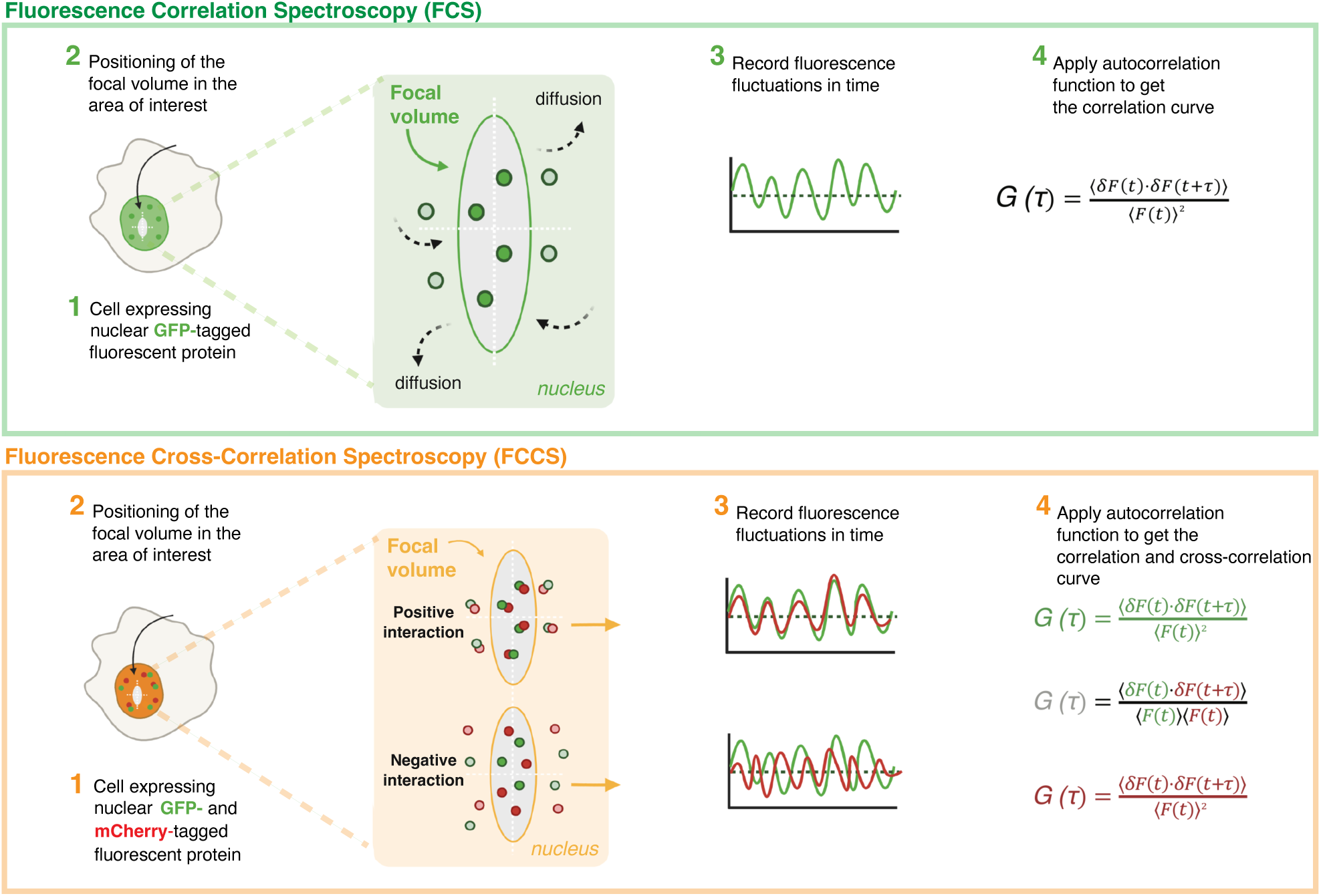
Schematic representation of fluorescence correlation spectroscopy (FCS) and fluorescence cross-correlation spectroscopy (FCCS) methods used in the study. Upper panel - FCS measurements are performed either in cells expressing a protein of interest fused to a GFP tag or in cell lysates from cells expressing the GFP fusion protein. Focal volume is positioned in a specific cellular location (live cell FCS) or in a fluorescent protein solution (cell lysate FCS). Fluorescence fluctuations are recorded and analysed by the autocorrelation function, which generates an autocorrelation curve, that is used to determine the diffusion time of the examined GFP-tagged protein. Lower panel - FCCS is based on the combined FCS measurements of two spectrally-distinct fluorescent proteins, in this case GFP and mCherry. Autocorrelation function is used for analysing diffusion of GFP- and mCherry-tagged proteins and also applied between channels, generating the cross-correlation curve. Interacting GFP- and mCherry-tagged proteins diffuse synchronously through the focal volume, so the cross-correlation curve (grey) is positive. In contrast, non-interacting GFP- and mCherry-tagged proteins diffuse independently from each other yielding a flat cross-correlation curve. The cross-correlation curves shown in the Figures reflect the interaction between the different fluorescently labelled molecules. The maximum possible correlation occurs when a molecule labeled with one fluorophore interacts with a molecule labelled with the other fluorophore. This interaction cannot happen more frequently than the presence of the most abundant molecule. Therefore, the FCCS curve can only reach a maximum value corresponding to the concentration of the most abundant molecule.

Based on the idea that pathogenic variants in structured protein domains disrupt macromolecular assemblies, we employed FCCS to rapidly assess *in vivo* protein complex formation and thereby functionally classify variants. We employed hereditary breast and ovarian cancer (HBOC) as a model disease because the molecular underpinning is well defined^12^. Moreover, data from a recent saturation genome editing (SGE) screening allowed us to compare our results with another functional test^4^. HBOC susceptibility genes are involved in homologous recombination repair (HRR), replication fork stability, DNA replication, and cell cycle checkpoint pathways. HRR is governed by a large assembly of factors, including BRCA1 and BRCA2, as well as PALB2, ATM, CHEK2, RAD51C, RAD51D, BARD1, RBBP8 and BRIP1. BRCA1 serves as scaffold for a number of these factors and pathogenic mutations are frequently found in the BRCA1 RING and BRCT domains that among other factors associate with BARD1 and RBBP8, respectively^13-15^. We show that FCCS, whether applied to full-length BRCA1 in live cells and/or to isolated domains in cellular lysates, reliably identify known BRCA1 RING or BRCT pathogenic variants. We also demonstrate the feasibility of employing the method for analysis of hereditary non-polyposis colorectal cancer (HNPCC)-related factor MSH2^16^ and MEN1 factor Menin^17^ in combination with DNA mismatch repair factor MSH6^18^ and transcription factor JUND^19^, respectively. Since the analysis can be completed in just a few hours, FCCS may be a useful complement to current clinical procedures for classifying genetic variants. Given its generic nature and design, we moreover envision that FCCS could serve as a valuable tool for variant classification in a wide variety of monogenic diseases.

## MATERIAL AND METHODS

### Cell lines

HeLa cells (ATCC® CCL-2™) were cultured in Dulbecco’s Modified Eagle Medium (DMEM), high glucose, no glutamine, no phenol red (Thermo Fisher Scientific, Cat. No. 31053028), supplemented with 1X GlutaMAX™ (Thermo Fisher Scientific, Cat. No. 35050038), 1 mM sodium pyruvate (Thermo Fisher Scientific, Cat. No. 11360039), 10% FBS (Biowest, Cat. No. BWSTS1810) and 1% penicillin/streptomycin (Thermo Fisher Scientific, Cat. No. 15070063) and were grown in a humidified incubator at 37 C and 5% CO_2_.

### Plasmids

The coding sequence of BRCA1 was PCR-amplified and cloned into pEGFP-C1 (Clontech) by restriction site digestion (SalI/SacII) and ligation. BARD1 and RBBP8 coding sequences were also PCR-amplified and cloned into pmCherry-C1 (Clontech). The sequences encoding for amino acids 1-109 and 1651-1864 of BRCA1 VCEP were separately cloned into pEGFP-C1 to generate vectors expressing the EGFP-tagged RING and BRCT trimmed domains, respectively. EGFP was PCR-amplified from pEGFP-C1 and cloned into pmCherry-C1 by restriction enzyme digestion in order to obtain the pmCherry-EGFP fusion protein. All inserts were confirmed by Sanger sequencing. Scale-up of plasmid DNA was performed with GeneJET Plasmid Midiprep Kit (Thermo Fisher Scientific, Cat. No. K0481).

### Cell seeding and plasmid transfection

For experiments in live cells, 200,000 HeLa cells were seeded in 4-well chambered glass-bottom coverslips (µ-Slide 4 Well Glass Bottom, Ibidi, Cat. No. 80427). 4-5 hours after, cells were transfected using FuGene HD transfection reagent (Promega, Cat. No. E2311). Briefly, 3 µl of FuGENE^®^ HD Transfection reagent was added to 50 µl of Opti-MEM^TM^ (Thermo Fisher Scientific, Cat. No. 31985062). Next, 500 ng of plasmid DNA were added, and the mixture was incubated for 15 minutes before addition to the wells. To achieve similar but slightly higher mCherry-tagged protein expression levels, GFP and mCherry vectors were mixed at a 2:1 ratio. For experiments in lysates, 250,000 cells were seeded per well in 6-well plastic-bottom plates (Nunc™ Cell-Culture Treated Multidishes, Thermo Fisher Scientific, Cat. No. 140675). 4-5 hours after, cells were transfected using FuGene HD transfection reagent. Briefly, 18 µl of FuGENE^®^ HD Transfection reagent were added to 600 µl of Opti-MEM^TM^. Next, 1 μg of plasmid DNA was added and the mixture was incubated for 15 minutes before 100 µl were added to each well. To achieve similar protein expression levels, GFP and mCherry vectors were mixed either at a 1:2 ratio (RING domain, RBBP8), 1:4 ratio (BRCT domain, RBBP8), 1:1 ratio (MSH6, MSH2), 1:1 (Menin, JUND).

### Confocal microscopy imaging

HeLa cells were seeded in 35 mm glass bottom dishes (No. 1.5 Coverslip 14mm G, uncoated, MatTek Corporation, Cat. No. P35G-1.5-14-C), transfected with GFP or/and mCherry vectors and subsequently imaged ∼24h after transfection. Confocal images were obtained using a Zeiss LSM780 confocal microscope with a Plan-Apochromat 63°ø/1.4 NA oil objective.

### Fluorescence Correlation Spectroscopy (FCS)

FCS measurements were recorded the day after transfection, selecting cells where the expression level of GFP and mCherry-tagged proteins was low. FCS measurements were performed with a Zeiss LSM780 confocal microscope using a C-Apochromat 40×/1.2 W Corr M27 objective and Immersion oil Immersol W 2010 (Zeiss). GFP or mCherry measurements were performed with an Argon laser with a 488 nm excitation wavelength (0.1% laser power) and a DPSS laser with a 561 nm excitation wavelength (0.1% laser power), respectively. GFP fluorescence was captured with a detection window of 482–553 nm and mCherry fluorescence was captured with a detection window of 590–695 nm, as described^20^. Before each measurement, average molecular count rate (kHz per molecule) was checked at different laser powers to ensure that fluorescence count signal was linear with laser power and not in saturation. Measurements were performed at randomly picked volumes in the cell nucleus, excluding nucleoli and other dense structures, for 1.5 minutes in 30 s intervals. The coverslip was taken from the incubator (37° C) and sealed with parafilm, before they were mounted on the microscope. The microscope measurements were taken at room temperature and coverslips were kept in the incubator until analyses. The experimental autocorrelation curves were obtained and analysed in ZEN 2011 software (Zeiss). The fits of the different models to the experimental data were also performed in ZEN 2011, using their in-built models as described ^20^.

### Fluorescence Cross-Correlation Spectroscopy (FCCS)

FCCS measurements in live cells were taken the day after transfection, selecting cells where the expression level of GFP and mCherry-tagged proteins was low. Recordings were performed in a Zeiss LSM780 confocal microscope using a C-Apochromat 40×/1.2 W Corr M27 objective and Immersion oil Immersol W 2010 (Zeiss). GFP or mCherry measurements were performed with an Argon laser with a 488 nm excitation wavelength (0.1% laser power) and a DPSS laser with a 561 nm excitation wavelength (0.1% laser power), respectively. GFP fluorescence was captured with a detection window of 482–553 nm and mCherry fluorescence was captured with a detection window of 590–695 nm. Before each measurement, average molecular count rate (kHz per molecule) was checked at different laser powers to ensure that fluorescence count signal was linear with laser power and not in saturation. Measurements were performed at randomly picked volumes in the cell nucleus for 1.5 minutes in 30 s intervals. As mentioned above nucleoli and other dense structures were excluded. The coverslip was taken from the incubator (37 ° C) and sealed with parafilm, and measurements of two wells (2/4) were taken 5–30 min later. Coverslip was placed back to the incubator for 30 min before taking the FCCS measurements of the remaining 2 wells. Experimental autocorrelation and cross-correlation curves were obtained and analysed in ZEN 2011 software (Zeiss). Cross-correlation and correlation amplitude values, needed to calculate the cross-correlation/correlation ratios, were extracted from the average of the amplitude values G(τ) in the interval from time points 8,00E-06 – 4,00E-04 from either curve in the area of maximum amplitude. Cross-correlation (CC)/autocorrelation (AC) ratio was calculated with the following formula: CC/AC ratio = [G(τ)CC – 1]/[G(τ)AC – 1].

FCCS on lysates was also performed the day after transfection. Cells were lysed at room temperature in 100 µl of lysis buffer containing 20 mM Tris-HCl pH 7.5, 140 mM KCl, 1.5 mM MgCl_2_, 1mM DTT and 0.5% NP-40 supplemented with 1:300 mammalian protease inhibitor cocktail (Sigma). Cell lysates were briefly centrifuged at 800 x g for 1 min and supernatant was transferred to a 35 mm glass bottom dish (No. 1.5 Coverslip 14mm G, uncoated, MatTek Corporation, Cat. No. P35G-1.5-14-C) before being subjected to FCCS. All the settings described above for FCCS analyses in live cells were also used for FCCS in lysates, except for the laser power, which was optimized as follows. For GFP-tagged RING domain variants and mCherry-tagged BARD1, GFP or mCherry measurements were performed with an Argon laser with a 488 nm excitation wavelength (0.3% laser power) and a DPSS laser with a 561 nm excitation wavelength (0.5% laser power), respectively. For GFP-tagged BRCT domain variants and mCherry-tagged RBBP8, GFP or mCherry measurements an Argon laser with a 488 nm excitation wavelength (0.4% laser power) and a DPSS laser with a 561 nm excitation wavelength (0.4% laser power) were used, respectively. The same laser settings were used for GFP-tagged MSH2 and mCherry-tagged MSH6 and GFP-tagged Menin and mCherry-tagged JUND.

## RESULTS

### Fluorescence Correlation Spectroscopy (FCS) of nuclear GFP-BRCA1

In order to assess whether FCS was able to portray the nuclear diffusion of BRCA1, HeLa cells were transiently transfected with a plasmid encoding GFP-BRCA1 or GFP for comparison. Since the autocorrelation is inversely correlated with the concentration of the fluorescent molecule under scrutiny, recordings were obtained from cells exhibiting low expression of the factors (Figure 2A). As illustrated by the normalized autocorrelation curves, nuclear GFP-BRCA1 diffuses about 8 times slower than GFP (Figure 2B) indicating that GFP-BRCA1, in contrast to GFP, is likely to be part of a larger assembly. In agreement with this, a subsequent fitting of the autocorrelation curve to models of 1, 2 or 3 components showed that the GFP-BRCA1 autocorrelation curve did not fit a 1-component diffusion model (blue line) (Figure 2, panels C and D). The poor fitting is illustrated by the residuals, which show the difference between the experimental data and the model fit. A better fit was observed after employing a free (3D) 2-component diffusion model (red line), although the fluctuation of the residuals was not completely random and fluctuating around zero. In contrast, fitting to a 3-component diffusion model (orange line) generated an almost random distribution of the residuals indicating that nuclear GFP-BRCA1 associates with a number of nuclear proteins.

**Figure 2.**
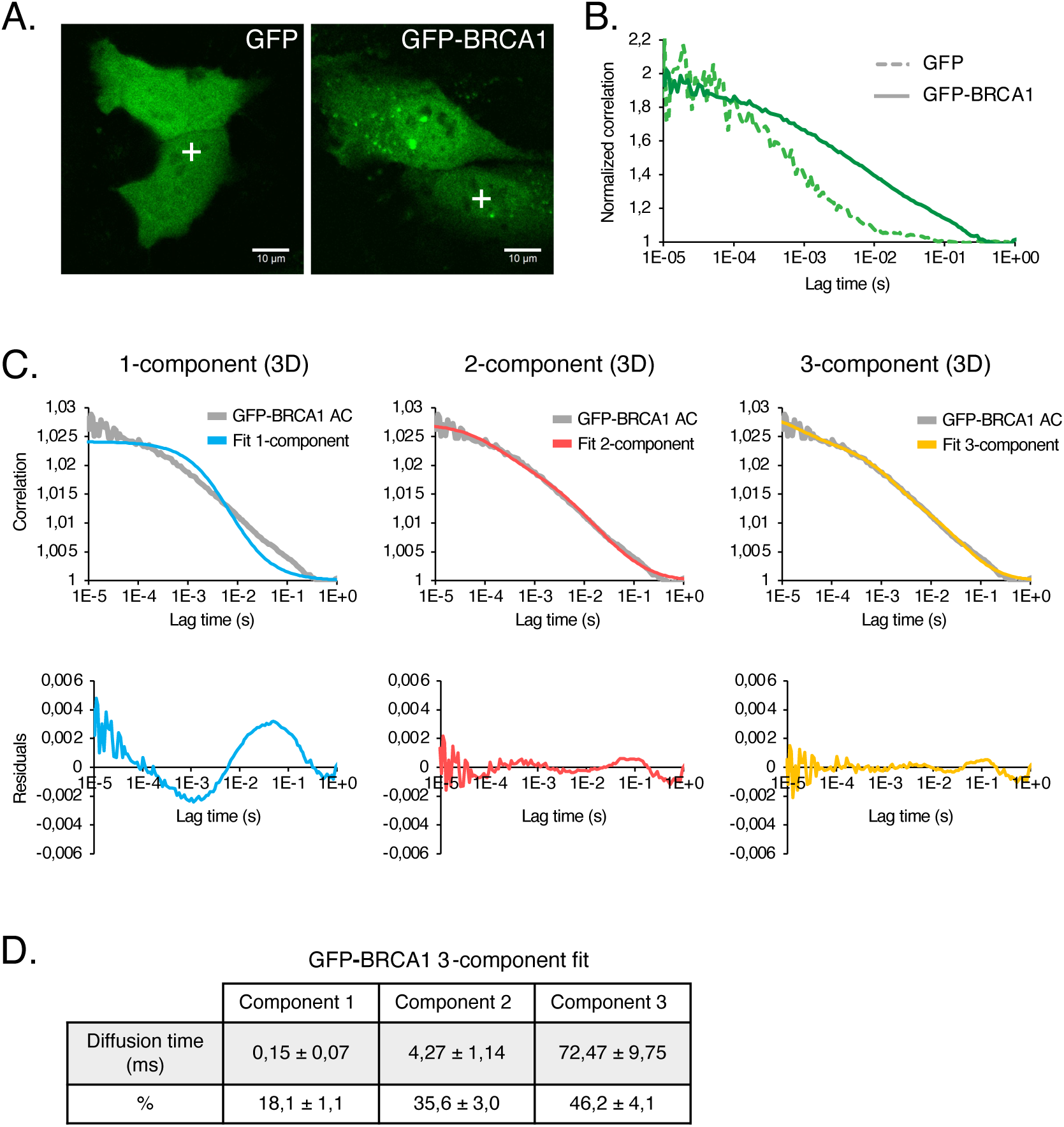
Complex formation of GFP-BRCA1 in live cells **A.** Confocal image of HeLa cells expressing GFP and GFP-tagged BRCA1. GFP exhibits a diffused fluorescent pattern throughout the cell while GFP-BRCA1 shows a prominent nuclear localization of the tagged protein. Crosses in the nuclear space show how focal volumes were arbitrarily positioned in the nucleoplasm to record the fluorescent measurements. **B.** Normalized autocorrelation curves for GFP and GFP-BRCA1 showing an averaged slower diffusion of GFP-BRCA1 compared to free GFP. **C.** Fitting of the GFP-BRCA1 autocorrelation curve to 1-, 2- and 3-component diffusion models (top) and their corresponding residuals (bottom). **D.** Diffusion time and percentage (%) of GFP-BRCA1 fitting to a 3-component diffusion model.

### Fluorescence Cross-Correlation Spectroscopy of BRCA1 binding to BARD1 and RBBP8

BRCA1 is an essential scaffold in the assembly of the HRR complex and the N-terminal RING domain of BRCA1 interacts with BARD1, whereas the C-terminal BRCT domain, among other factors, associates with RBBP8 (CtIP) ^13-15^. We therefore exploited the possibility to detect and quantify complex formation with these two factors by Fluorescence Cross-Correlation Spectroscopy (FCCS). Cells were co-transfected with plasmids encoding GFP-BRCA1 and mCherry-BARD1 or mCherry-RBBP8, respectively. For comparison, we compared the results to a GFP-mCherry fusion construct^20^ (Supplemental Figure 1). Nuclear FCCS measurements were performed in the nucleoplasm avoiding the nucleolus, matrix and dense foci with conceivably immobile GFP-BRCA1. For each measurement, the GFP-BRCA1 and mCherry-BARD1 or mCherry-RBBP8 autocorrelation curves and the cross-correlation curve were obtained. Measurements were routinely obtained from 5-10 different cells and in each cell, we performed at least 3 recordings of 30 s. Unstable correlation curves distorted by bleaching or instability of the stage were omitted from the analysis. We also favoured cells exhibiting low levels of the expressed factors in order to reach correlations above 1.005, although this was not always feasible. GFP-BRCA1, mCherry-BARD1 and mCherry-RBBP8 are mainly nuclear, although a significant fraction of BRCA1 and BARD1 also remains in the cytoplasm. In cells exhibiting nuclear speckles the factors co-existed indicating they were part of the same complexes (Figure 3, panels A and B). Figure 3, panels C and D, shows the averaged correlation curves for GFP-BRCA1, mCherry-BARD1 or mCherry-RBBP8 and the corresponding cross-correlation, respectively. The averaged cross-correlation for mCherry-BARD1 and -RBBP8 were 28% (STDEV 8%) and 36% (STDEV 11%), respectively. For comparison the mCherry-GFP fusion protein exhibited a cross-correlation of 33% (STDEV 5%) (Supplemental Figure 1). Panel D shows the variation of the correlation amplitudes among different cells. At the described inter-replicate STDEV, the assay may detect a reduced cross-correlation of >35% with a *P* value equal to or less than 0.05. Since the correlation readings obtained in live cells are relatively low and variable compared to measurements in solutions (see below), we averaged correlation values over a series of 20 readings (time points 8.00E-06 – 4.00E-04 seconds) in order to establish the plateau of the correlation curves as indicated in panels C and D. Only replicates exhibiting a STDEV of <0.01 among the plateau values were used for the analysis so readings exhibiting high variation at the plateau were excluded.

**Figure 3.**
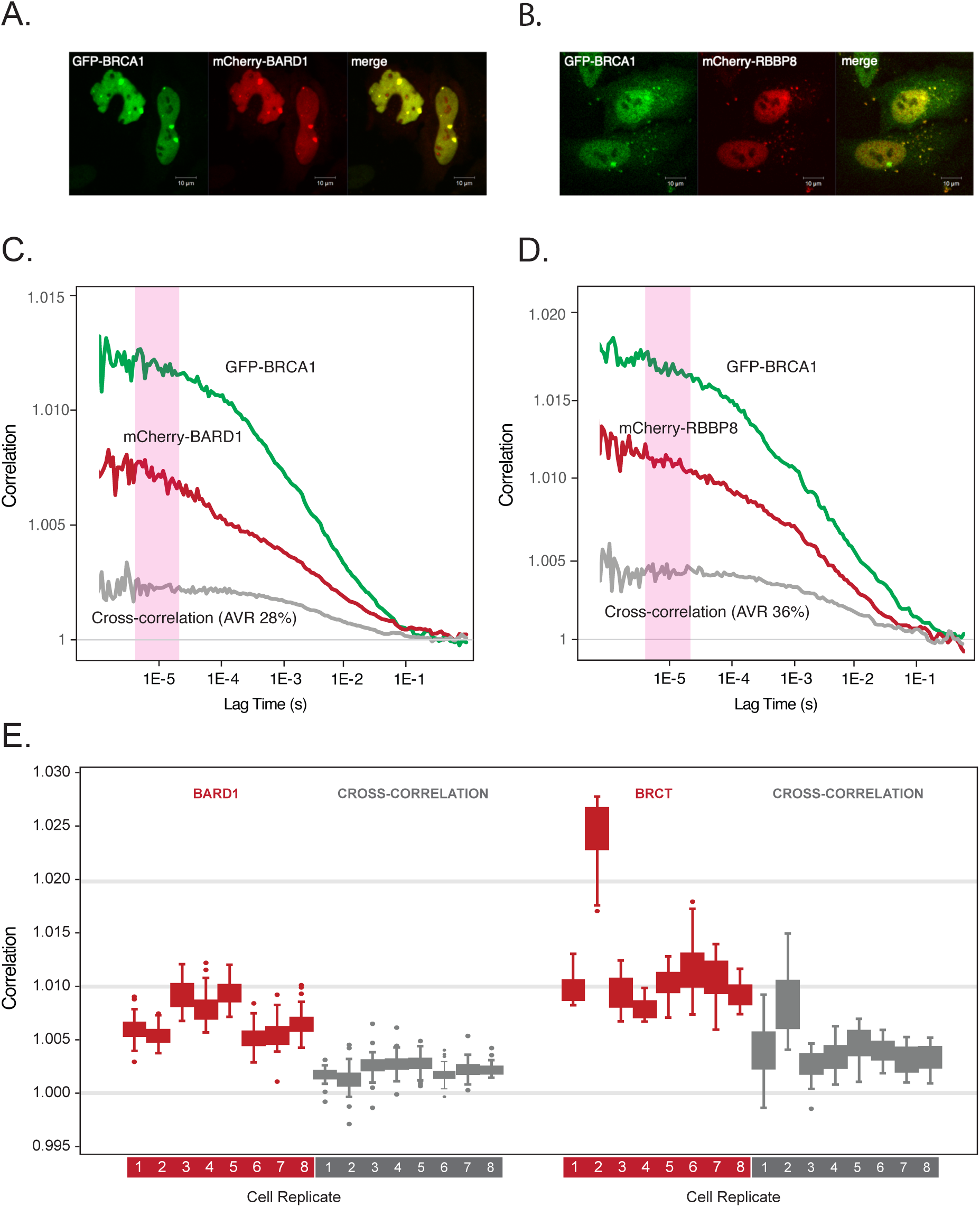
Association of BRCA1 with BARD1 and RBBP8/CtIP in live cells. **A.** Confocal images of HeLa cells expressing GFP-BRCA1 (left) and mCherry-BARD1 (middle), together with the merged (right) image. **B.** Confocal images of HeLa cells expressing GFP-BRCA1 (left) and mCherry-RBBP8/CtIP (middle), together with the merged (right) image. **C.** Autocorrelation and cross-correlation curves obtained from GFP-BRCA1 and mCherry-BARD1 nuclear measurements. **D.** Autocorrelation and cross-correlation curves obtained from GFP-BRCA1 and mCherry-RBBP8 nuclear measurements. The pink columns in C and D show the range of autocorrelation values that were averaged to determine the plateau of the curves. **E.** The chart shows the distribution and variation among and within the absolute averaged autocorrelation values for the BARD1 and RBBP8 plateau and their corresponding cross-correlation curves in 8 different cells (labelled 1-8), respectively.

### FCCS analyses of BRCA1 RING and BRCT domain variants in live cells

We subsequently examined binding of a series of known benign (ACMG 1 and 2), VUS (ACMG 3) and pathogenic variants (ACMG 4 and 5) in the BRCA1 RING and BRCT domains, respectively. The predicted AlphaFold structure ^21^ of the two domains in the context of the entire BRCA1 is depicted in Figure 4 and the position of the included variants is outlined in the blow-up of the two domains. The RING and BRCT domains are positioned in the core of BRCA1 surrounded by what are presumably large stretches of intrinsically disordered regions. Variants were selected so they covered the domains and represented a distribution of benign and pathogenic variants. Moreover, they should be registred in ClinVar or in the recent SGE analyses^4^. The RING module is considered to extend from amino acids 2 to 101 (BRCA1 VECP), which includes two flanking alpha helixes governing heterodimerization with BARD1 (amino acids 2-23 and 65-101), as well as, the catalytic RING domain (amino acids 24-64). The BRCA1 BRCT domain is composed of two tandem BRCT modules from amino acids 1650-1857 (BRCA1 VECP). A few benign variants located in immediate proximity to the BRCA1 RING (Tyr105Cys) and BRCT (Pro1859Arg) domains were also included.

**Figure 4.**
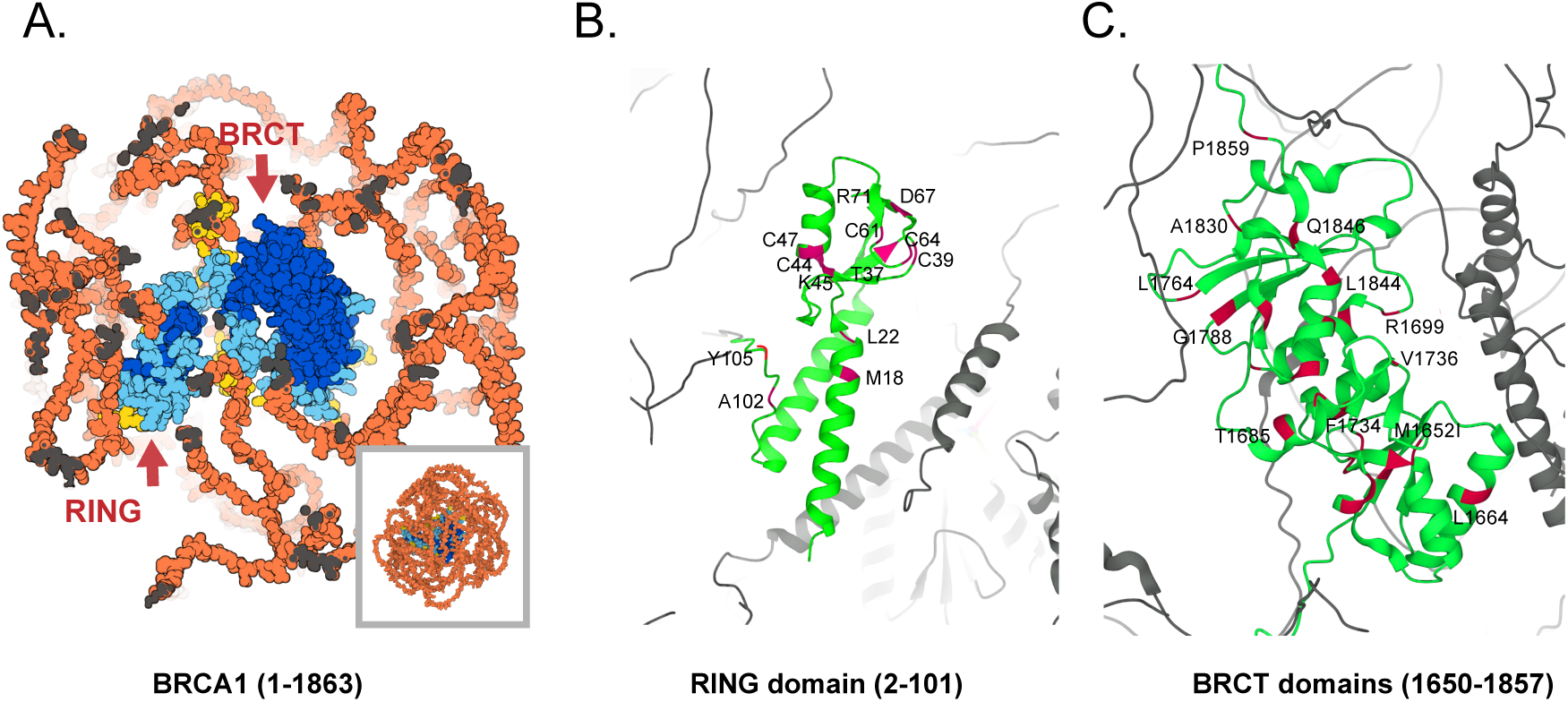
**A.** Space filling representation of the predicted AlphaFold structure of BRCA1 (UniProt identifier P38398). The amino acids are coloured according to the AlphaFold per-residue model confidence score (pLDDT) from very high (pLDDT > 90) (dark blue), high (90 > pLDDT > 70) (light blue) to low (70 > pLDDT > 50) (yellow) and very low (pLDDT < 50) (orange). The conserved RING and BRCT domains are located in the core of the protein and embedded by presumably unstructured stretches. **B.** Shows a ribbon of the RING domain composed of the BARD1-interacting alpha helices (lower part) and the Zn^2+^ coordinated ligase (upper part). The position of the examined variants is shown in red together with the corresponding amino acid. **C.** Shows a ribbon of the two BRCT domains with the tested variants labelled in red. The position of selected amino acids is shown to facilitate identification of the labelled variants.

Figure 5, panels A and B, shows the averaged correlation curves of wild-type BRCA1 in combination with two RING or BRCT variants exhibiting reduced cross-correlation. The results from all tested variants are shown below in panels C and D. The distribution and variation of the correlation values in the plateau for the individual replicates for the variants are shown in Supplemental Figure 2. With the exception of T37K and R71G variants, the pathogenic RING mutations reduced cross-correlation between GFP-BRCA1 and mCherry-BARD1 (*P*<0.02), while the benign mutations had no significant effect on the binding (Figure 5, panel C and Table 1). C44F reduced binding ∼50%, whereas the observed reduction of remaining pathogenic variants ranged from 35%-45% of the wild-type. The pathogenic R71G substitution is considered to affect mRNA splicing^22,23^ (Table 1) and was not expected to display impaired binding between BRCA1-BARD1 in our assay. T37K, in contrast, locates to a pocket in the proximity of the RING ligase and was examined in more detail employing isolated domains (see below). We also tested the M18T variant but were unable to express the protein at a sufficient level for analysis. We surmised that the mutant was unstable and this variant was also examined in the context of isolated domain. Pathogenic variants in the BRCT module clearly reduced RBBP8 binding, and the binding data were concordant with both ClinVar classification, SGE screening and AlphaMissense (Figure 5, panels C and Table 1). Variants such as M1775R, G1788V and R1699W, strongly reduced RBBP8 binding and were almost indistinguishable from the truncating L726fs and K991* controls. A number of the benign BRCT variants were also found to reduce binding and as shown in Figure 5, panel C, the variants appear to represent a continuum of reduced binding. To distinguish pathogenic BRCT variants the significance level should consequently be adjusted 0.001 and the binding to less than 40% of the wild type. T1684A is clearly a borderline variant that although classified as benign exhibits an almost 50% reduction (*P*<0.002) in binding. Table 1 delineates the results from the FCCS analysis and shows the corresponding classifications from ClinVar, SGE and AlphaMissense.

**Figure 5.**
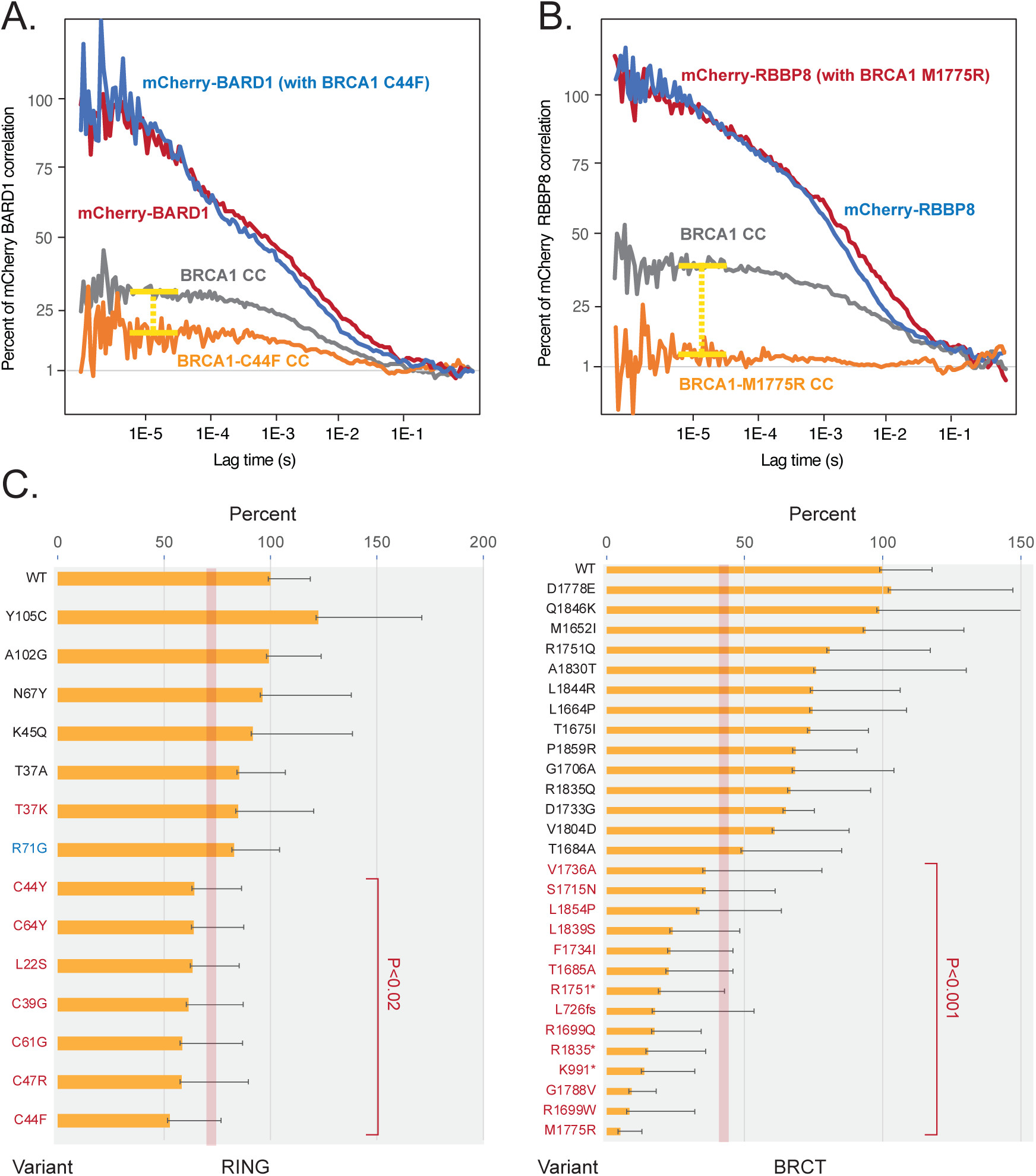
Impaired binding of BRCA1 variants to BARD1 and RBBP8/CtIP in live cells. **A.** Normalized mCherry-BARD1 correlation and cross-correlation curves with GFP-BRCA1 wild-type or GFP-BRCA1-C44F (curves not shown). **B.** Normalized mCherry-RBBP8 autocorrelation and cross-correlation curves with GFP-BRCA1 wild-type and GFP-BRCA1 M1775R (curves not shown). **C.** Averaged and normalized cross-correlation values of RING variants and BRCT variants, respectively. Recordings were obtained from 5-10 different cells and in each cell, we performed at least 3 recordings of 30 s. The columns are ordered according to the reduction in binding. Benign variants or VUS are labelled in black, whereas, pathogenic variants are labelled in red. The R71G splice variant is labelled in blue. The error-bars represents the STDEV and the *P* values (*t-*test, two-tailed, unequal variance) are indicated.

**Table 1.** Summary of the classification of the examined BRCA1 RING and BRCT variants by FCCS, ClinVar, saturation genomic editing (SGE)^4^ and AlphaMissense^2^. The location, frequency, rsIDs and SGE scores are indicated for each variant. *M18T and T37R results only from lysate analysis with isolated RING domain.

| Transcript | Protein | FCCS | ClinVar | SGE | SGE score | SGE RNA | AlphaMissense | Frequency | rs ID |
| --- | --- | --- | --- | --- | --- | --- | --- | --- | --- |
| **RING** |  |  |  |  |  |  |  |  |  |
| c.53T>C | p.Met18Thr* | LOB | 5 | LOF | -1.89 | -1.1 | Likely pathogenic | 0 | rs80356929 |
| c.65T>C | p.Leu22Ser | LOB | 5 | LOF | -2.11 | -0.8 | Likely pathogenic | 0 | rs80357438 |
| c.109A>G | p.Thr37Ala | Binding | 4/5 | Functional | 0.11 | 0.1 | Likely pathogenic | NA | rs2055253295 |
| c.110C>A | p.Thr37Lys* | AMB | 5 | LOF | -2.49 | -0.8 | Likely pathogenic | 0 | rs80356880 |
| c.110C>G | p.Thr37Arg* | LOB | 4/5 | LOF | -2.49 | -0.7 | Likely pathogenic | 0 | rs80356880 |
| c.115T>G | p.Cys39Gly | LOB | 4/5 | LOF | -2.22 | -0.6 | Likely pathogenic | 0 | rs80357164 |
| c.131G>A | p.Cys44Tyr | LOB | 5 | LOF | -2.40 | 0.0 | Likely pathogenic | NA | rs80357446 |
| c.131G>T | p.Cys44Phe | LOB | 5 | LOF | -2.54 | -0.9 | Likely pathogenic | NA | rs80357446 |
| c.133A>C | p.Lys45Gln | Binding | 1 | Functional | -0.70 | -0.1 | Ambiguous | 0.00003 | rs769650474 |
| c.139T>C | p.Cys47Arg | LOB | 4/5 | LOF | -1.84 | -0.5 | Likely pathogenic | 0 | rs80357370 |
| c.181T>G | p.Cys61Gly | LOB | 5 | LOF | -1.74 | -0.3 | Likely pathogenic | 0 | rs28897672 |
| c.191G>A | p.Cys64Tyr | LOB | 5 | LOF | -2.05 | 0.1 | Likely pathogenic | 0 | rs55851803 |
| c.199G>T | p.Asp67Tyr* | AMB | 1 | Functional | -0.08 | 0.0 | Likely benign | 0.00001 | rs80357102 |
| c.211A>G | p.Arg71Gly | Binding | 5 | LOF | -1.49 | -2.8 | Likely pathogenic | 0 | rs80357382 |
| c.305C>G | p.Ala102Gly | Binding | 1 | NA | - | NA | Likely benign | 0.00001 | rs80357190 |
| c.314A>G | p.Tyr105Cys | Binding | 1 | NA | - | NA | Likely benign | 0.00002 | rs28897673 |
| **BRCT** |  |  |  |  |  |  |  |  |  |
| c.2176_2177del | Leu726fs | LOB | 5 | NA | - | NA | NA | NA | rs397508945 |
| c.2971A>T | p.Lys991* | LOB | 5 | NA | - | NA | NA | NA | rs886040090 |
| c.4956G>A | p.Met1652Ile | Binding | 1 | Functional | 0.14 | -0.4 | Ambiguous | 0.01118 | rs1799967 |
| c.4991T>C | p.Leu1664Pro | Binding | 1 | Functional | -0.01 | -0.2 | Likely benign | 0 | rs80357314 |
| c.5024C>T | p.Thr1675Ile | Binding | 1 | Functional | 0.04 | 0.0 | Likely benign | 0 | rs150729791 |
| c.5050A>G | p.Thr1684Ala | AMB | 3 | Functional | -0.65 | 0.1 | Ambiguous | NA | rs879255491 |
| c.5053A>G | p.Thr1685Ala | LOB | 5 | LOF | -1.84 | -0.3 | Ambiguous | NA | rs80356890 |
| c.5096G>A | p.Arg1699Gln | LOB | 5 | LOF | -2.21 | 0.0 | Likely pathogenic | 0.00002 | rs41293459 |
| c.5095C>T | p.Arg1699Trp | LOB | 5 | NA | - | NA | Likely pathogenic | 0 | rs55770810 |
| c.5117G>C | p.Gly1706Ala | Binding | 1 | Functional | -0.05 | -0.2 | Ambiguous | 0.00008 | rs80356860 |
| c.5144G>A | p.Ser1715Asp | LOB | 5 | LOF | -2.29 | -0.5 | Likely pathogenic | 0 | rs45444999 |
| c.5198A>G | p.Asp1733Gly | Binding | 1 | Functional | 0.17 | 0.2 | Likely benign | 0.00003 | rs80357270 |
| c.5200T>A | p.Phe1734Ile | LOB | 3 | LOF | -2.26 | -0.6 | Likely pathogenic | NA | rs80356957 |
| c.5207T>C | p.Val1736Ala | LOB | 5 | LOF | -1.60 | 0.2 | Likely pathogenic | 0.00003 | rs45553935 |
| c.5252G>A | p.Arg1751Gln | Binding | 1 | Functional | -0.15 | -0.2 | Likely benign | 0.00007 | rs80357442 |
| c.5251C>T | p.Arg1751* | LOB | 5 | LOF | -1.88 | -0.1 | NA | 0 | rs80357123 |
| c.5291T>C | p.Leu1764Pro | LOB | 5 | LOF | -1.99 | 0.4 | Likely pathogenic | NA | rs80357281 |
| c.5324T>G | p.Met1775Arg | LOB | 5 | LOF | -1.39 | -0.6 | Likely pathogenic | 0 | rs41293463 |
| c.5334T>A | p.Asp1778Glu | Binding | 3 | Functional | -0.09 | -0.2 | Likely benign | 0 | rs754152768 |
| c.5363G>T | p.Gly1788Val | LOB | 5 | LOF | -1.68 | -0.4 | Likely pathogenic | 0 | rs80357069 |
| c.5411T>A | p.Val1804Asp | Binding | 1 | Functional | 0.43 | 0.0 | Likely benign | 0.00001 | rs80356920 |
| c.5488G>A | p.Ala1830Thr | Binding | 3 | Functional | 0.17 | -0.2 | Likely benign | 0 | rs80357393 |
| c.5503C>T | p.Arg1835* | LOB | 5 | LOF | -2.30 | -1.4 | NA | 0 | rs41293465 |
| c.5516T>C | p.Leu1839Ser | LOB | 5 | LOF | -2.49 | -0.7 | Likely pathogenic | NA | rs398122702 |
| c.5531T>G | p.Leu1844Arg | Binding | 1 | Functional | -0.06 | -1.2 | Likely benign | 0.00007 | rs80357323 |
| c.5536C>A | p.Gln1846Lys | Binding | 3 | Functional | -0.09 | -0.1 | Likely benign | 0 | rs80356873 |
| c.5561T>C | p.Leu1854Pro | LOB | 3 | LOF | -1.34 | -0.9 | Likely pathogenic | NA | rs80356996 |
| c.5576C>G | p.Pro1859Arg | Binding | 1 | NA | - | NA | Likely benign | 0.0001 | rs80357322 |

### FCCS analyses of isolated BRCA1 RING and BRCT domain variants in cell lysates

We recently developed an FCCS protocol employing cell lysates to complement live cell readings^20^. Lysates do not reconcile the spatial resolution of live cells, but they have less spatial constraints and provide optimal readings because they can be diluted, which allows you to discern even small changes in binding. Moreover, in contrast to the single live cell analysis, the assay is a simple and reproducible bulk analysis providing less variation between replicates. Briefly, cells were transfected with the trimmed BRCA1 GFP-RING or GFP-BRCT domains in combination with mCherry-BARD1 or mCherry-RBBP8, respectively, followed by iso-osmolar lysis in buffer containing non-ionic detergent to release the cytoplasmic content. Measurements, were performed on a drop of lysate positioned at the bottom of the same type of glass dishes that were used for live recordings. For each variant, we performed 12 readings in 3 biological replicates. Correlations ranged from 1.5-2.2, which is about 50 times higher than in live cells. The averaged results from the analysis of selected variants are shown in Figure 6C and the associated replicate data is shown in Supplemental Figure 3, panels B and C. For the wild-type RING domain, cross-correlation was 25% of the mCherry-BARD1 correlation and for the BRCT domain the cross-correlation with mCherry-RBBP8 was 17%. The isolated domains reconciled to a large extent the results from the full-length BRCA1, and as illustrated in Figure 6A, the higher accuracy of the assay was able to depict small differences in binding of the T37K, T37R and T37A variants of the RING domain. We detected a minor but significant (*P*<0.05) reduction in the binding of pathogenic T37K variant and the T37R variants decreased binding with 30%. We classify T37K as ambiguous and the T37R as pathogenic. Also, the binding of D67Y was impaired by ∼22%. Moreover, unlike live cells, the M18T variant in the isolated domain was adequately expressed and found to exhibit reduced binding classifying it as damaging. For the BRCT domain, we were also able to identify pathogenic variants in concordance with the live cell analysis (Figure 6B and 6C). Compared to the RING domain, BRCT variants in general had slightly more severe effects on binding and similar to live cells even benign variants appeared to cause a small reduction in binding to RBBP8. Taken together, we conclude that isolated structural modules may serve as a convenient and sensitive way to classify genetic variants.

**Figure 6.**
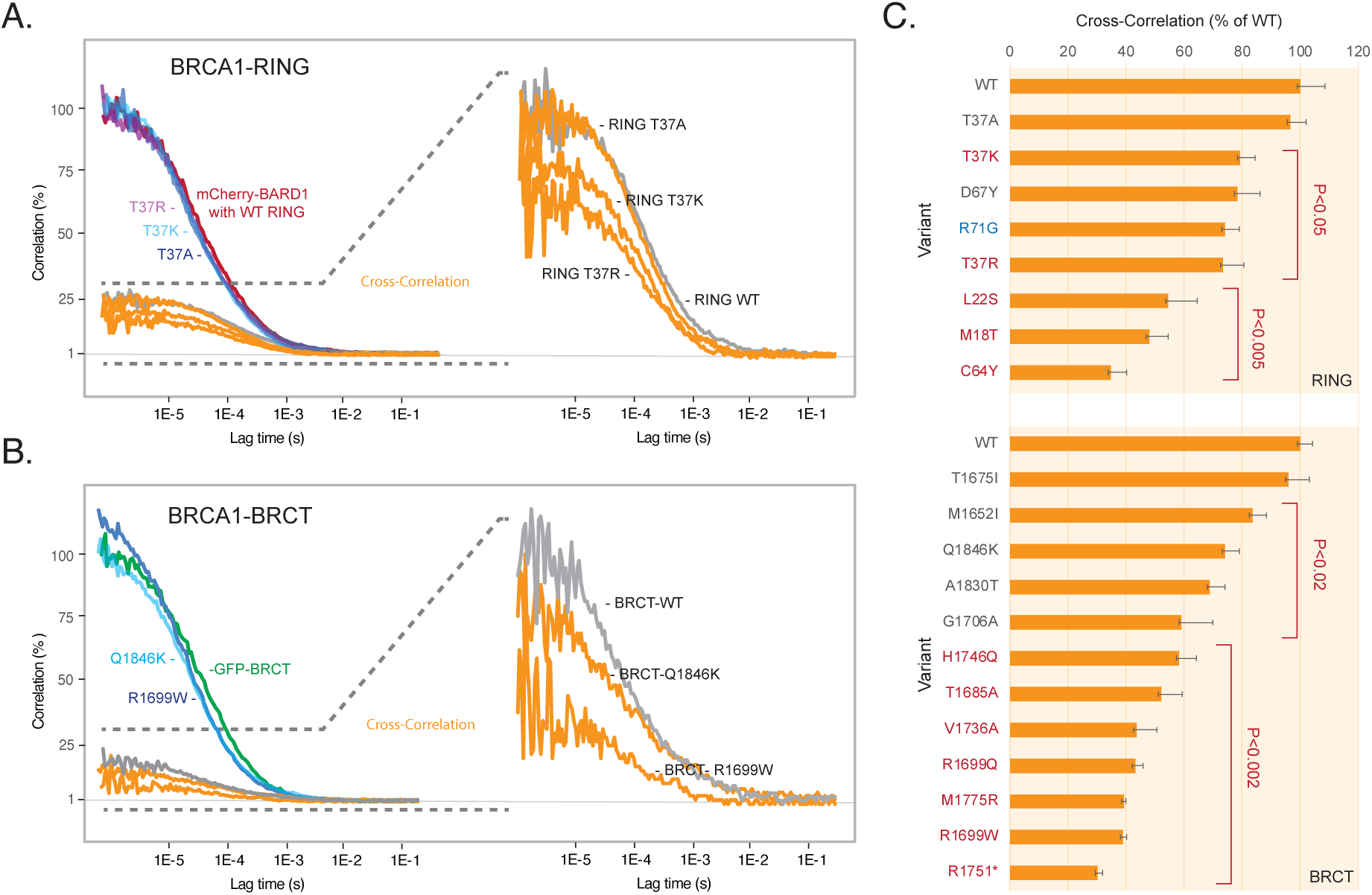
BRCA1 variant analysis employing isolated RING and BRCT domains in cell lysates. **A.** Cells were transfected for 24 h with GFP-RING or with GFP-RING-T37A, GFP-RING-T37K or GFP-RING-T37R in combination with mCherry BARD1 before they were solubilized and the lysates examined by FCCS. The panel shows the normalized mCherry-BARD1 correlation curve (red) and corresponding wild-type RING cross correlation (grey) as well as the mCherry-BARD1 correlation and cross-correlation curves (orange) from the analyses of the GFP-tagged RING-T37A (dark blue), T37K (light blue) or T37R (purple) variants. The right panel is a blow up of the cross-correlation curves from each variant. **B.** Correlation curves of GFP-BRCT (green) and mCherry-RBBP8/CtIP (red) and the corresponding wild-type cross-correlation curve (grey) as well as the GFP-BRCT autocorrelation (blue) and cross-correlation curves (orange) from the analyses of the GFP-tagged BRCT-Q1846K and R1699W variants. **C.** Normalized averaged cross-correlation values from RING and BRCT variants. The columns are ordered according to the reduction in binding. Benign variants or VUS are labelled in black, whereas, pathogenic variants are labelled in red. The R71G splice variant is labelled in blue. The results are derived from three independent experiments and error bars indicate the STDEV. The error-bars represents the STDEV and the *P* values (*t-*test, two-tailed, unequal variance) are indicated. The intra-assay variations shown in Supplemental Figure 3.

### Application of FCCS analyses to MSH2-MSH6 and Menin-JUND heterodimers

To explore if FCCS could be applied to other structural domains, we performed assays with MSH2 and MSH6, that are involved in hereditary non-polyposis colorectal cancer (HNPCC), and Menin, that is encoded by the multiple endocrine neoplasia (*MEN1*) gene, in combination with JUND. The analyses were performed as described for BRCA1, and in live cells, we observed a strong cross-correlation of 34% between MSH2 and MSH6 in both the nucleus and in the cytoplasm (Figure 7A). Cross-correlation was significantly reduced by insertion of pathogenic variants MSH2 P622L and C697F^9,24^ (Figure 7A). In lysates cross-correlation was slightly lower (27%), but as in live cells the two pathogenic variants caused a significant reduction in cross-correlation. In lysates, we could moreover observe a left shift in the correlation curves, indicating that the variants are likely to be excluded from an even larger protein assembly. The interaction between Menin and JUND was examined in lysates and this revealed a cross-correlation of 21%. Two variants - A237V and A242V^25^, that in ClinVar were categorized as VUS and likely pathogenic, respectively, and according to AlphaMissense are likely pathogenic, reduced binding to approximately 50% of the wild-type. Taken together, we infer that FCCS is feasible for variant classification in various structural domains.

**Figure 7.**
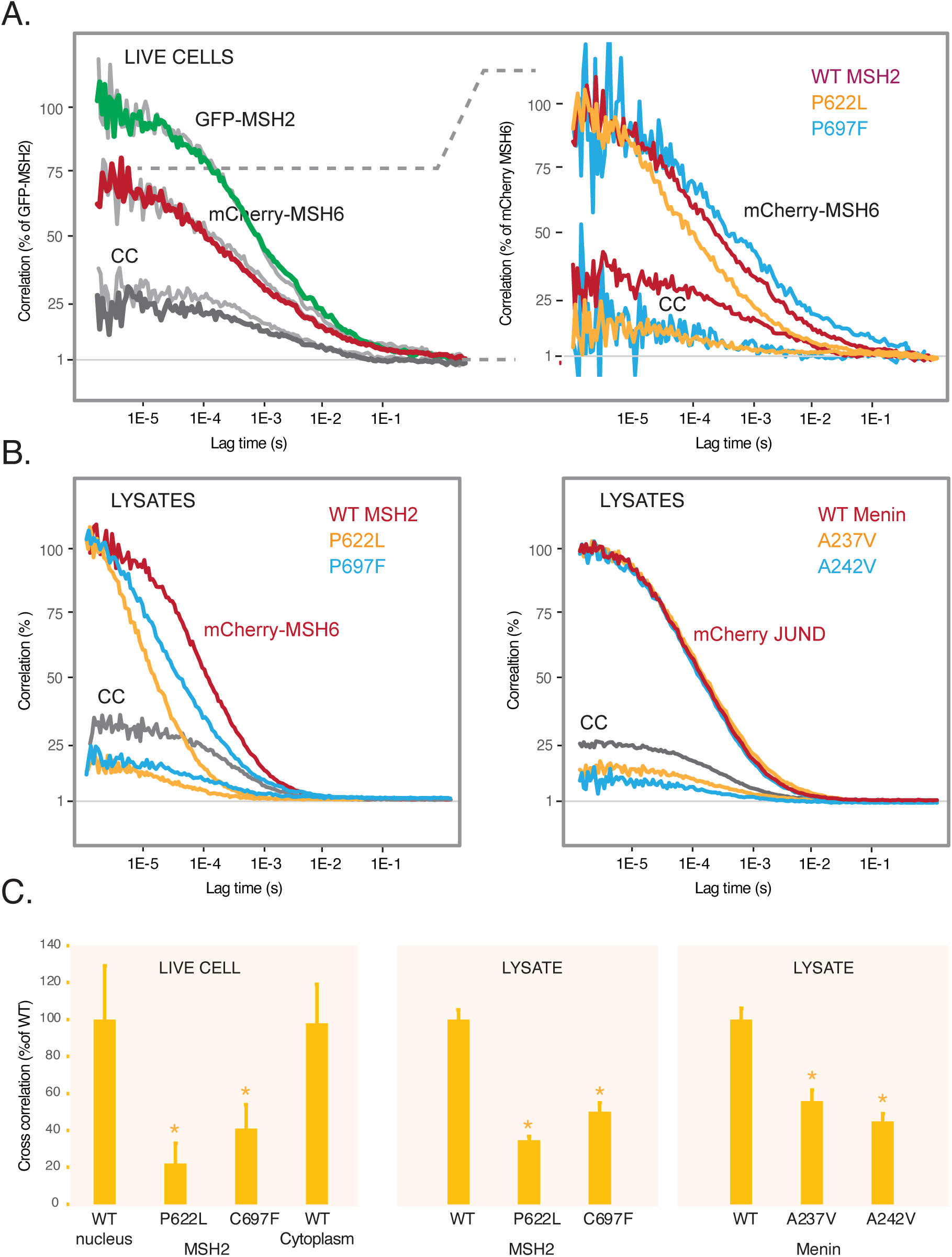
FCCS analyses of MSH2 and Menin. **A.** Nuclear correlation (green and red) and cross-correlation curves (dark grey) from cells transfected with GFP-MSH2 and mCherry-MSH6. The light grey curves show the same analyses where the focal volume was positioned in the cytoplasm. The blow up to the right shows the analyses of two pathogenic variants P622L and C697F. **B.** FCCS analyses of the same MSH2 constructs in cellular lysates (left) and with GFP-Menin and mCherry-JUND (right) or with JUND in combination with Menin A237V (NM_001370259.2(MEN1):c.710C>T) or A242V (NM_001370259.2(MEN1):c.725C>T) variants (same as A242V, A247V - MEN1_HUMAN ENST00000337652.5). **C.** Normalized cross-correlation results from three independent experiments. Error bars represents the STDEV and the asterisks indicate *P*<0.05 *(t*-test, two-tailed, unequal variance).

## DISCUSSION

Key cellular processes, including transcription, mRNA processing and transport, protein synthesis, signalling events, and metabolism, hinge upon the orchestration of protein assemblies with varying degrees of complexity. While protein complexes may exhibit homomeric structures, more frequently they involve an interplay of distinct proteins. Binding is typically governed by conserved domains characterized by well-defined structures, and less commonly by intrinsically disordered regions^26,27^. Many monogenic diseases are associated with variants that disrupt the formation of specific functional assemblies^28-30^, e.g. hereditary cancer syndromes, developmental abnormalities resulting from disruptions in cytoskeletal or ciliary architecture, as well as metabolic, mitochondrial diseases, and late-onset conditions like ALS^31^. Recent data in fact suggest that pathogenic mutations are concentrated at protein interfaces, underscoring the potential utility of analysing protein interactions to characterize the pathogenic significance of missense variants^26^. Generally, missense variants impact protein structure by inducing misfolding occasionally leading to the degradation of the involved factors. Based on the idea that analysis of complex formation could represent a sensitive and generic way of identifying pathogenic variants, we employed fluorescence correlation and cross-correlation spectroscopy to rapidly and precisely assess *in vivo* protein complex formation.

The limitations of FCS and FCCS are mainly regarding fluorophore tagging that may affect the folding and bioactivity of some proteins. BRCA1 can be tagged without major loss of biological activity and GFP-BRCA1 has previously been employed in the characterisation of its role in double-strand break (DSB) sites^32^. In agreement with earlier studies, our initial characterisation of the GFP-BRCA1 construct showed that it located to characteristic nuclear foci and complexed with other proteins. Association between BRCA1 and BARD1 and RBBP8/CtIP was close to 30%, which is similar to that obtained with a GFP-mCherry fusion protein, showing that binding is regular and stable (Supplemental Figure 1). Due to photobleaching, misfolding or fluorescent proteins being “off” or in dark states^33^, cross-correlation is not expected to be 100%. In our experience it is useful to separate the protein under investigation and the fluorophore by a short flexible linker sequence^20^. We have moreover experienced that the relative abundance of the factors should be as similar as possible and care should be taken to work in the linear range of the photodetector. Transient expression systems provide relatively high concentrations of the expressed proteins so cells exhibiting low levels of expression should in general be chosen for analysis. Whenever possible and meaningful, we prefer the lysate-based analysis since the concentration of the factors can be accurately controlled to achieve the best possible correlation amplitude and accuracy of the assay. Moreover, lysates are a bulk analysis which reduces the pronounced cell to cell variation, that we observed for some variants in the single cell live analysis. Finally, it is important to underscore that the assay can only be considered for its positive (reduced binding) predictive value since protein binding in principle may occur flawlessly despite loss of e.g. enzymatic activity or incorrect splicing as described below.

In general, the FCCS findings aligned with the current SGE, ClinVar, and AlphaMissense classifications of the examined variants. Reduced binding was associated with pathogenic or ambiguous variants in both live cells and in isolated domains in lysates. Two known pathogenic variants, T37K and R71G, exhibited normal binding in live cells. The solvent exposed RING T37 locates to a small cavity between the BRCA1-BARD1 heterodimerization interface and the catalytic site^34^. There are three known variants at this position (T37K, T37R and T37A) and the results provide an example of the limitations of the live cells analysis. Only in lysates using the isolated RING domain, it was apparent that T37K had a small but significant effect on binding, whereas the T37R exhibited a greater loss of binding, likely reflecting that arginine, which forms a larger number of electrostatic interactions, perturbed the overall structure to a larger extent. R71G has as mentioned been documented to reduce splicing^22^ and the reduced mRNA expression is also reflected in the SGE analysis (Table 1). Since the employed constructs are intron-less it was expected to be similar to the wild-type, although we noted a small reduction in binding. Finally, D67Y also exhibited a small reduction in binding in lysates and although generally classified as benign some reports have in fact suggested that D67Y may be hypomorphic ^35-37^, we have therefore labelled the variant as ambiguous. For BRCT variants, we observed a single discrepancy with the SGE in the case of the T1684A variant, that reduce binding moderately by ∼50%, whereas the SGE shows a reduced activity (score -0.62) that was insufficient to classify the variant as pathogenic. The difference may be explained by the thresholds of the assays and in agreement with a small but significant functional reduction, ClinVar depicts the variant as a VUS and AlphaMissense find it is ambiguous.

The domain-specific analyses reconciled the results from live cells but also revealed that variants overall had a slightly larger impact on binding in the isolated domains. This was particularly evident for variants in the conserved BRCT domains, where even benign variants reduced binding to RBBP8. In fact sorting of the variants based on their binding shows that variants represents a continuum from wild-type activity to severe reduction in binding. Pathogenic variants were clearly more severe than the benign and could be identified by a combination of reduction in binding and significance level. The specific cut-off level obviously needs to be established between known pathogenic and benign variants during clinical implementation for other monogenic diseases. The impact of variants classified as likely benign underscores the biological significance of larger molecular assemblies, in the sense that the concerted action of many factors may stabilise binding of each individual factor. Moreover, in the specific case of BRCA1 the results could point at a role for the long stretches of intrinsically disordered sequence. The results also indicate that deleterious variants in the future maybe are better qualified by quantitative measures rather than a binary (likely)benign – (likely)pathogenic way. An example is the T1684A variant that reduce binding to a level not very different from known pathogenic variants and therefore could be envisioned to be harmful for susceptible persons. Recent data have indicated that protein-folding chaperones may overcome the pathogenicity of variants in the BRCA1 C-terminal (BRCT) domain. HSP70 binds pathogenic BRCA1-BRCT variants and the magnitude of binding was correlated to loss of folding and function^38^. If the chaperone activity varies in the population some individuals may be particularly susceptible to variants that otherwise are classified as benign. Chemical chaperones have moreover successfully been applied to re-establish the function of mutant proteins in monogenic diseases^38-41^ and in this scenario quantitative methods such as FCCS may be helpful to identify relevant variants and monitor the effect of the chaperone.

We finally examined the feasibility of employing FCCS for analysis of other domains and variants in MSH2 and Menin, respectively. Since both proteins are significantly smaller than BRCA1 the live analysis were considerably easier to perform because the transfection efficiency was many fold higher. MSH2 is composed of five structural domains including a DNA mismatch binding domain, a connector and a lever domain with an incorporated Clamp domain and a C-terminal ATPase domain^42^. The pathogenic P622L and C697F variants are positioned in the ATPase domain in two distinct beta strands. MSH2 and MSH6 exhibited a high cross-correlation (37%) and binding was clearly reduced by the two variants. Interestingly, we also observed that impaired heterodimerization leads to a significant increase in the diffusion constant of cytoplasmic MSH2, suggesting that other factors may be part of the complex even at this stage. Menin is a single domain scaffold protein regulating gene transcription and cell signalling^43^. The protein has been described to attain the shape of a curved hand and JUND binds the central pocket spanning residues 27–47, which is relatively far from the mutated R237 and R242. The observed decrease in binding serves to underscore that the entire amino acid composition of this type of proteins may have been fine-tuned during evolution and support the concept of structural based analysis for classification of genetic variants.

Taken together FCCS is reproducible and fast and only requires a confocal microscope and lab space for cell transfection. Including the time for transfection and recordings, the analyses can be completed in just two workdays, making FCCS appealing for the clinical environment. We do not envision FCCS for systematic functional screens like those conducted for BRCA1^4^ or MSH2^44^ variants, but the technology hold promise for the functional assessment of individual patient variants in a wide range of monogenic diseases, where there is no available functional test. We examined four different protein interactions involving various structured domains and successfully identified pathogenic variants in all of them. Thus, FCCS may serve as a potential generic approach for functional variant assessment which could provide additional information to the current ACMG classification scheme.

## Data Availability

All data produced in the present study are available upon reasonable request to the authors

## ACKNOWLEDGEMENTS

Lelde Kalnina is thanked for help with the generation of the MSH2 variants.

