## Supplemental Figures for "Functional Assessment of Protein Variants in Structured Domains by Fluorescence Cross-Correlation Spectroscopy"

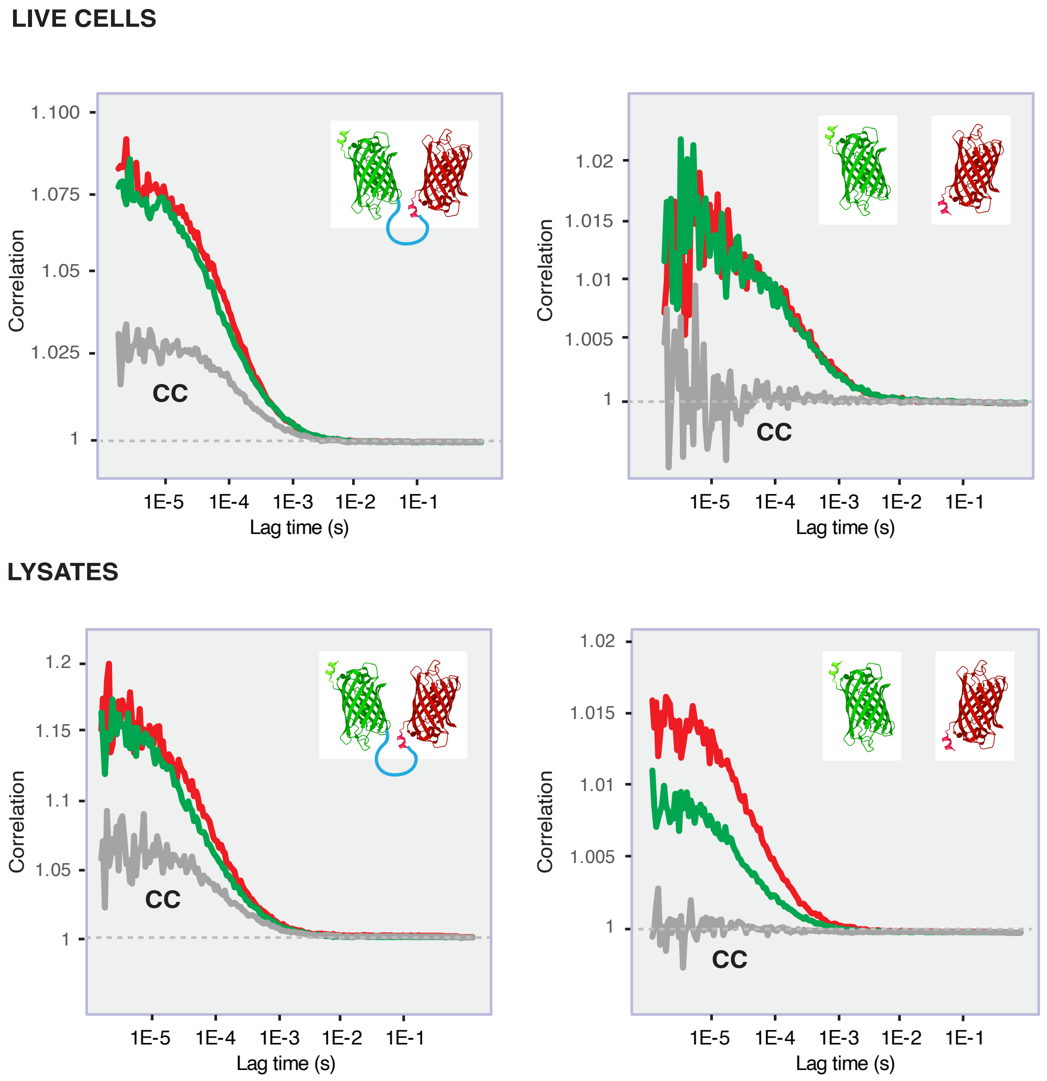


**Supplemental Figure 1.** Cross-correlation of EGFP and mCherry fusion proteins.
HeLa cells were transfected with either separate EGFP-C1 and mCherry-C1 vectors (right panel) or with a EGFP-mCherry fusion construct (left panel) (20) before FCCS recordings were performed in live cells (upper panels) or in lysates (lower panels). Both in live cells and lysates we observed a cross-correlation of ~35% in cells transfected with the fusion protein, whereas no cross-correlation was observed in the cells transfected with separate EGFP and mCherry vectors.
Cross-correlation is not expected to reach 100% since a fraction of the molecules will undergo photo-bleaching and quenching, moreover some fluorescent proteins may be ‘off’ or in dark states.

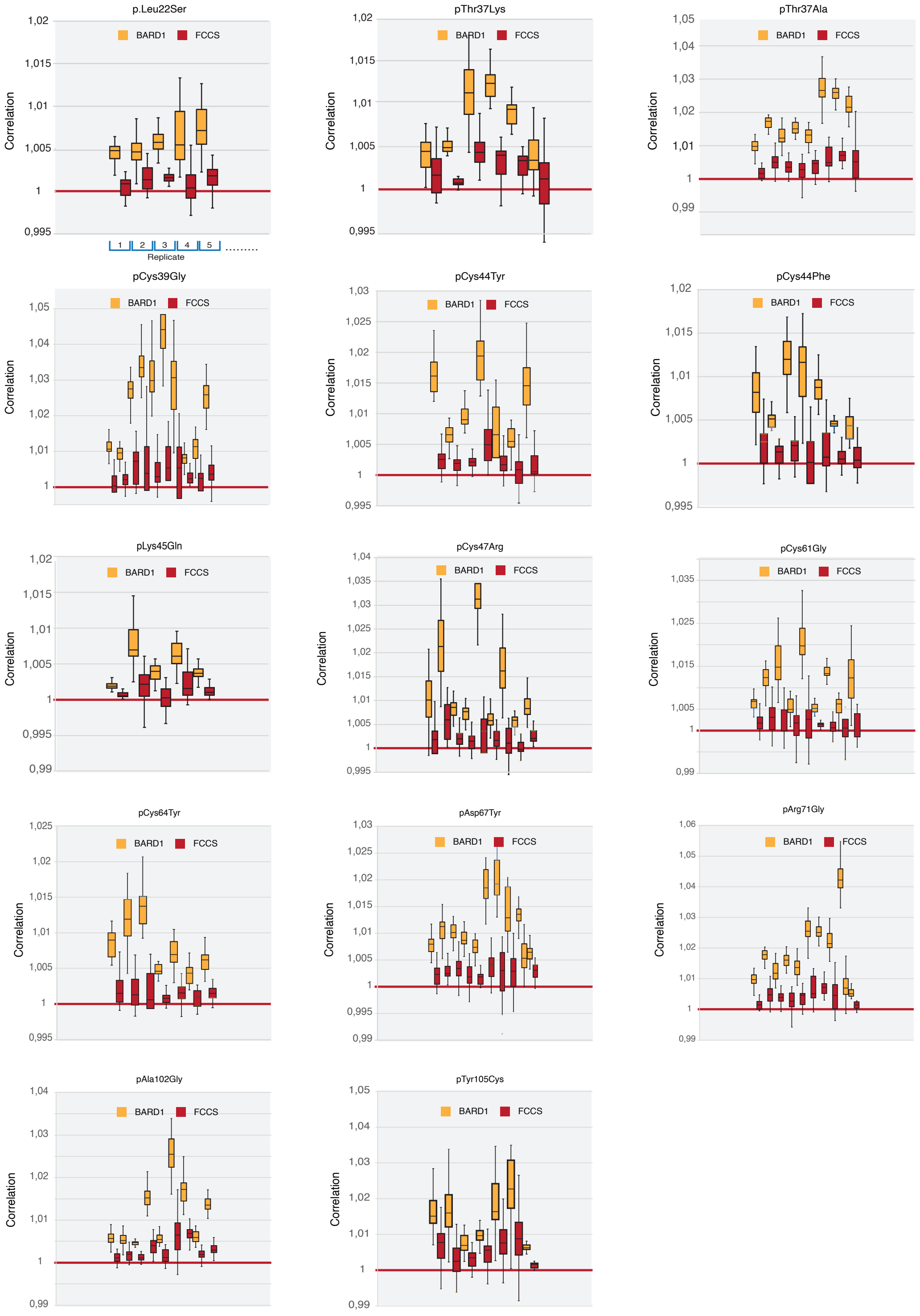
p.Leu22Ser
pThr37Lys
pThr37Ala
pCys39Gly
pCys44Tyr
pCys44Phe
pLys45Gln
pCys47Arg
pCys61Gly
pCys64Tyr
pAsp67Tyr
pArg71Gly
pAla102Gly
pTyr105Cys
BARD1
FCCS
Correlation
Replicate
1
2
3
4
5

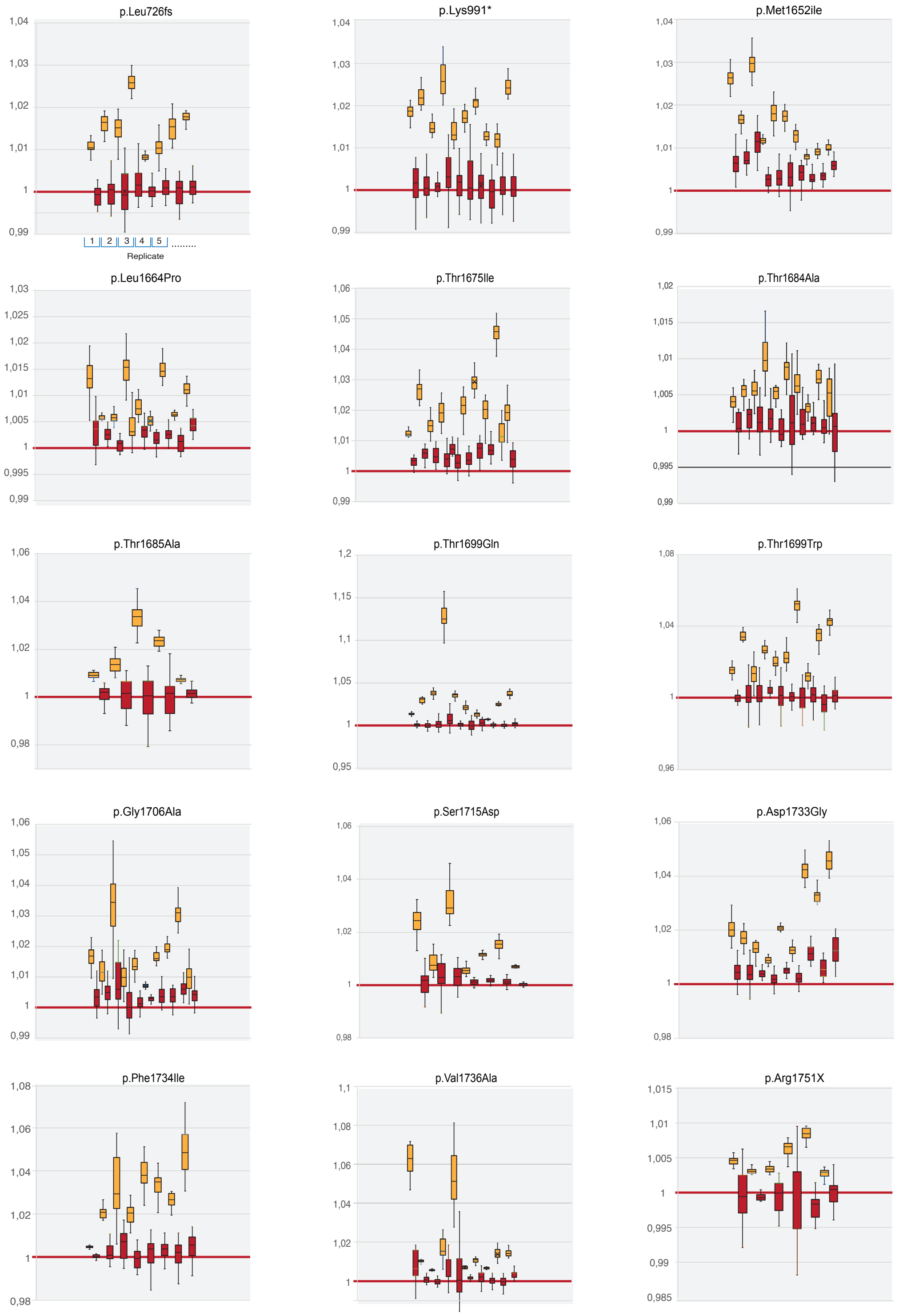

p.Leu726fs
1,04
1,03
1,02
1,01
1
0,99
1 2 3 4 5 .........
Replicate
p.Lys991*
1,04
1,03
1,02
1,01
1
0,99
p.Met1652ile
1,04
1,03
1,02
1,01
1
0,99
p.Leu1664Pro
1,03
1,025
1,02
1,015
1,01
1,005
1
0,995
0,99
p.Thr1675Ile
1,06
1,05
1,04
1,03
1,02
1,01
1
0,99
p.Thr1684Ala
1,02
1,015
1,01
1,005
1
0,995
0,99
p.Thr1685Ala
1,06
1,04
1,02
1
0,98
p.Thr1699Gln
1,2
1,15
1,1
1,05
1
0,95
p.Thr1699Trp
1,08
1,04
1
0,96
p.Gly1706Ala
1,06
1,05
1,04
1,03
1,02
1,01
1
0,99
p.Ser1715Asp
1,06
1,04
1,02
1
0,98
p.Asp1733Gly
1,06
1,04
1,02
1
0,98
p.Phe1734Ile
1,08
1,06
1,04
1,02
1
0,98
p.Val1736Ala
1,1
1,08
1,06
1,04
1,02
1
p.Arg1751X
1,015
1,01
1,005
1
0,995
0,99
0,985

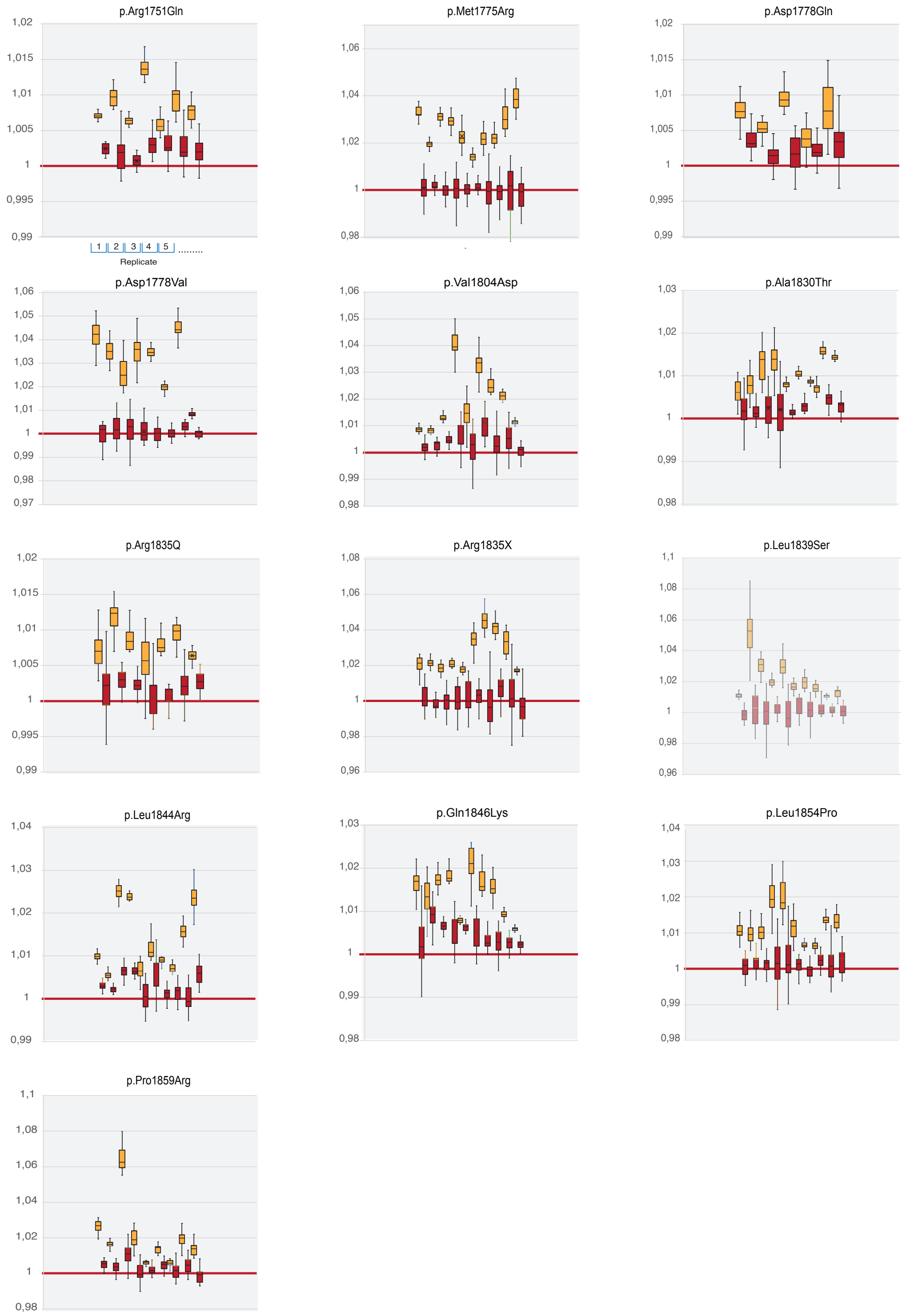


**Supplemental Figure 2.** Overview of correlation and cross correlation data from the BRCA1 variant analysis. The charts show for each tested variant the distribution and variation of the 20 correlation values from the time points 8,00E-06 – 4,00E-04. BARD1 and RBBP8 FCS correlation values are filled with orange and the corresponding FCCS values are marked in red. The individual readings raging from 5-10 replicates (cells) are shown next to each other as indicated in the first chart.

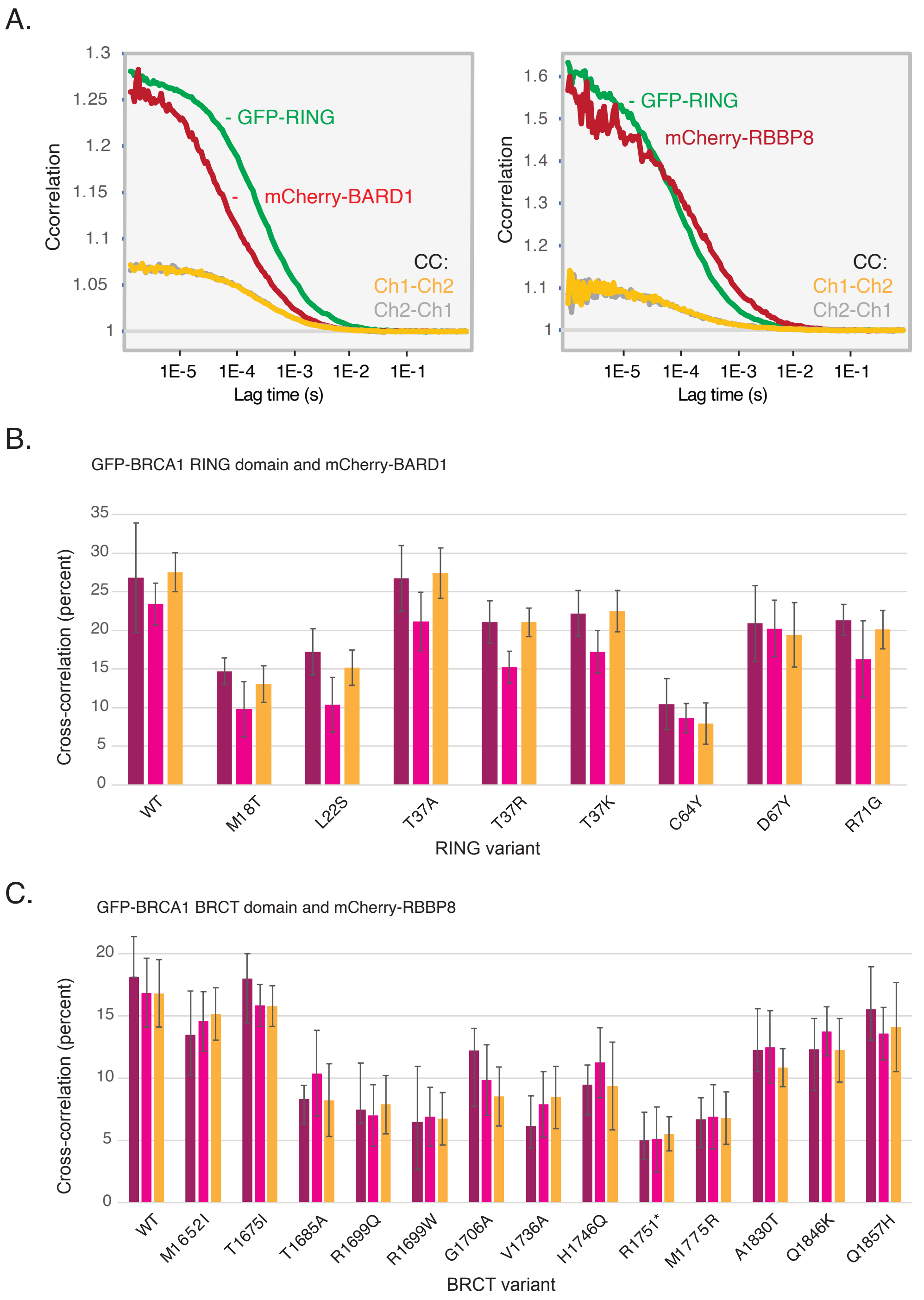


**Supplemental Figure 3**

**A.** Representative averaged correlation curve from analyses of cell lysates from cells transfected with RING (left panel) or BRCT (right panel) domains, respectively. Correlations are indicated in absolute values. **B. and C.** Inter-assay variation of RING or BRCT domain FCCS in cellular lysates. The columns show the percent of cross-correlation from three independent biological replicates. Each replicate comprise 12 averaged recordings. The SDEV represents the variation among the individual recordings. Each recording takes about 10 minutes and including cell transfection and lysis the assay of a variant may be completed over 2 days. Q1857 is located just outside of the BRCT and was included as control.
